# Trend and Factors Associated with Cardiovascular Risk in Peru: An Analysis of the National Demographic and Family Health Survey, 2014-2022

**DOI:** 10.1101/2024.05.22.24307711

**Authors:** Víctor Juan Vera-Ponce, Fiorella E. Zuzunaga-Montoya, Luisa Erika Milagros Vásquez-Romero, Joan A. Loayza-Castro, Mario J. Valladares-Garrido, Carmen Inés Gutierrez De Carrillo

## Abstract

**Introduction:** Cardiovascular diseases (CVD), which encompass a broad array of disorders impacting both the heart and circulatory system, remain a predominant source of mortality and morbidity that takes a toll on populations around the world.

**Objective:** This study focuses on the prevalence and trends of cardiovascular risk (CVR) in Peru from 2014 to 2022 and the factors associated with these conditions.

**Methods:** We conducted a cross-sectional analytical study using a database from the National Survey of Demography and Family Health collected from 2014 to 2022, covering individuals aged 40 to 75. Measurements included blood pressure, smoking, and body mass index (BMI), among others, using the World Health Organization’s (WHO) risk calculator. Stratified sampling methods were applied, and participants lacking interest variables were excluded.

**Results:** The prevalence of low and high CVR was 74.89% and 4.48%, respectively. Variations in CVR trends were observed from 2014 to 2022. Factors significantly associated with higher CVR included male sex, older age, higher education, wealth, smoking habits, obesity, and health conditions such as hypertension and type 2 diabetes.

**Conclusions:** The study revealed a significant prevalence of CVR factors in the studied population. The critical importance of addressing these prevalent factors in public health policies and strategies for preventing and managing cardiovascular diseases is strongly highlighted.

## Introduction

Cardiovascular diseases (CVD), which encompass a broad array of disorders impacting both the heart and circulatory system, remain a predominant source of mortality and morbidity that takes a toll on populations around the world. Due to the widespread nature and significance of public wellness of various cardiovascular events, several risk assessment tools have been created to gauge an individual’s potential likelihood of future cardiovascular occurrences. These tools are essential for identifying at-risk persons and implementing appropriate preventive strategies (1,2).

Globally, these diseases sadly claim nearly 18 million lives annually, comprising over 30 percent of the total lives lost around the planet each year (3). CVD is the leading cause of death in nations such as the United States, impacting huge numbers of individuals and exerting considerable strain on medical infrastructure. Within the context of Latin America, cardiovascular diseases are significantly challenged as epidemiological transition, lifestyle changes, and the widespread existence of risk factors like hypertension and diabetes drive their prevalence upwards (4,5). Recent trends in Peru have seen a growth of cardiovascular risk (CVR) elements, which have contributed to a growing strain from these ailments afflicting the population. While research into this area has begun, there remains a notable lack of targeted studies and accessible statistics regarding the circumstances facing people in Peru (6).

Given this scenario, it is imperative to investigate CVR in Peru thoroughly. Determining the prevalence and trends of CVR through national surveys, such as the one conducted from 2014 to 2022, and analyzing the associated factors becomes a crucial tool for formulating effective health policies. This study seeks to fill this gap, providing essential data for designing preventive and intervention strategies tailored to the Peruvian reality.

## Material and Methods

### Design

We conducted a cross-sectional analytical study using a database from the National Survey of Demography and Family Health (ENDES) collected from 2014 to 2022 (7). To ensure a detailed report of observational studies in the field of epidemiology, our study adhered to the STROBE guidelines (8).

### Population, Eligibility Criteria, and Sample

The survey was conducted nationwide and included Peruvians aged 15 to 99 years from urban and rural areas across 24 departments in Peru. ENDES implemented a two-stage stratified sampling method, which was probabilistically independent for rural and urban areas. For technical details, refer to ENDES technical documents (7). Our study included individuals aged 40 to 75, as the CVR method is designed for these age ranges. Those without the variables of interest were excluded.

### Assessment of Consistency and Plausibility of Measurements

Parameters reflecting the methodology of previous related analyses were established to ensure only practical blood pressure readings were included in the studies. This required that Systolic Blood Pressure (SBP) and Diastolic Blood Pressure (DBP) measurements stay within established limits: SBP readings between 70 mmHg and 270 mmHg, and DBP between 50 mmHg and 150 mmHg. Measurements not meeting these plausibility criteria were excluded from further analysis (9).

### Variables and Measurement

The primary variable was the calculation of CVR. For this, we decided to use the WHO risk calculator (10), as the database used does not have laboratory analyses that would be necessary for other such instruments. Variables used for this calculation were age (in years), systolic blood pressure (SBP), smoking presence, and body mass index (BMI). Subsequently, the CVR score was categorized into low risk (<10%), moderate risk (between 10% and <20%), and high risk (≥20%). However, CVR was dichotomized for regression analysis into low/moderate and high risk.

To define the presence of smoking, the question assessed: Do you currently use any tobacco products like cigarettes, cigars, or pipes? (Yes vs No). Self-reported information on diabetes mellitus type 2 (T2DM) history was evaluated by a question also coded as no versus yes: Has a doctor or other health worker ever told you that you have high blood sugar levels or diabetes? (Yes vs No).

The research considered various demographic and health factors, including gender (female or male), age range (20 to 35, 36 to 59, 60 to 69, and over 70 years), marital status (married or single), geographical location (Metropolitan Lima, other coastal areas, highlands, or jungle), socioeconomic level (from poorest to most affluent), an education level (no education or primary education versus secondary education or higher), place of residence (urban or rural), presence of physical disability (yes or no), daily consumption of more than five servings of fruits and vegetables (yes or no), existence of type 2 diabetes mellitus (T2DM) (yes or no), altitude levels (0 to 499, 500 to 1499, 1500 to 2999, 3000 or more), racial identification (Other, Quechua, Aymara, Native or Amazonian Indigenous, Black, White/Caucasian or Mestizo), nutritional status, abdominal obesity (yes or no), smoking habit, and alcohol consumption (11).

Racial self-identification was determined through self-report, asking participants if they considered belonging to any of the mentioned groups based on their ancestors and customs.

For the evaluation of abdominal obesity, criteria from the Adult Treatment Panel III of the Cholesterol Education Program (WC-ATPIII) were followed, defining a WC ≥ 102 cm for men and a waist ≥ 88 cm for women (12). The WHO classification was used to determine nutritional status based on BMI, establishing the following cutoff points for adults: average weight (< 25 kg/m^2), overweight (BMI between 25 and 29.9 kg/m^2), and obesity (BMI ≥ 30 kg/m^2).

Participants’ smoking habits were determined through their reports, categorizing them into non-smokers, former smokers, or active smokers (13). Similarly, alcohol consumption was based on the self-assessment of the interviewees and divided into categories: never consumed or not consumed in the last 12 months; moderate consumption (≥ once in the previous 30 days, but <5 drinks in the last year in men or <4 in women) and excessive consumption (≥ once in the last 30 days and ≥ five drinks in men or ≥ four in women) (14).

### Procedures

A formalized process for gauging measurements uniformly was instituted to ensure consistent and dependable determinations of systolic and diastolic blood pressure. A digital sphygmomanometer (OMRON, model HEM-713) was used to record the blood pressure of participants, using cuffs of two sizes according to each person’s arm circumference: standard (220–320 mm) and large (330–430 mm).

Measurements were taken in a controlled environment to ensure participants’ comfort, seated with the right arm at heart level on a flat surface. Having rested for five minutes, the initial measurement was obtained, with a second determination coming two minutes afterward to follow the first. This interval allowed for the stabilization of cardiovascular parameters. After that, the mean of the systolic and diastolic blood pressure readings for each participant was derived, with this average value then serving as a point of reference throughout the ensuing analytical processes. This method helped stabilize variations in blood pressure and provided a more accurate picture of the individual’s usual blood pressure level, offering clinicians a more reliable assessment tool.

### Statistical Analysis

The statistical program R version 4.03 was used to obtain the results. This analysis was divided into three tables and one graph.

The first table was descriptive, summarizing the data in absolute and relative frequencies. The second was a bivariate analysis to see if there were differences between the covariables and CVR; for this, Rao & Scott’s chi-square was used, as it must be considered that the sampling is complex. For the third table, adjusted prevalence ratios (PRa) were calculated through a generalized linear model of the Poisson family with Robust variance.

Finally, a trend graph was made between the years 2014 and 2022 to see if there were changes over the years.

### Ethical Considerations

This study used anonymous and publicly accessible data, which do not include information that could personally identify participants, thus eliminating any ethical risk to them. The annual data collection of ENDES is carried out by the National Institute of Statistics and Informatics of Peru. This government entity ensures obtaining informed consent from participants for data collection through the survey; consent is collected directly for participants over 18 years old. In the case of minors under 18, consent is requested from their parents or legal guardians, thus ensuring ethics in the evaluation of minors.

## Results

### Participants

Table 1 presents a percentage breakdown of a population’s various sociodemographic and health variables. It details the proportion of women, 48.24%, and divides age into three groups, with those aged 70 and over encompassing 7.46%. Regarding educational level, 29.18% have higher education.

**Table 1.**
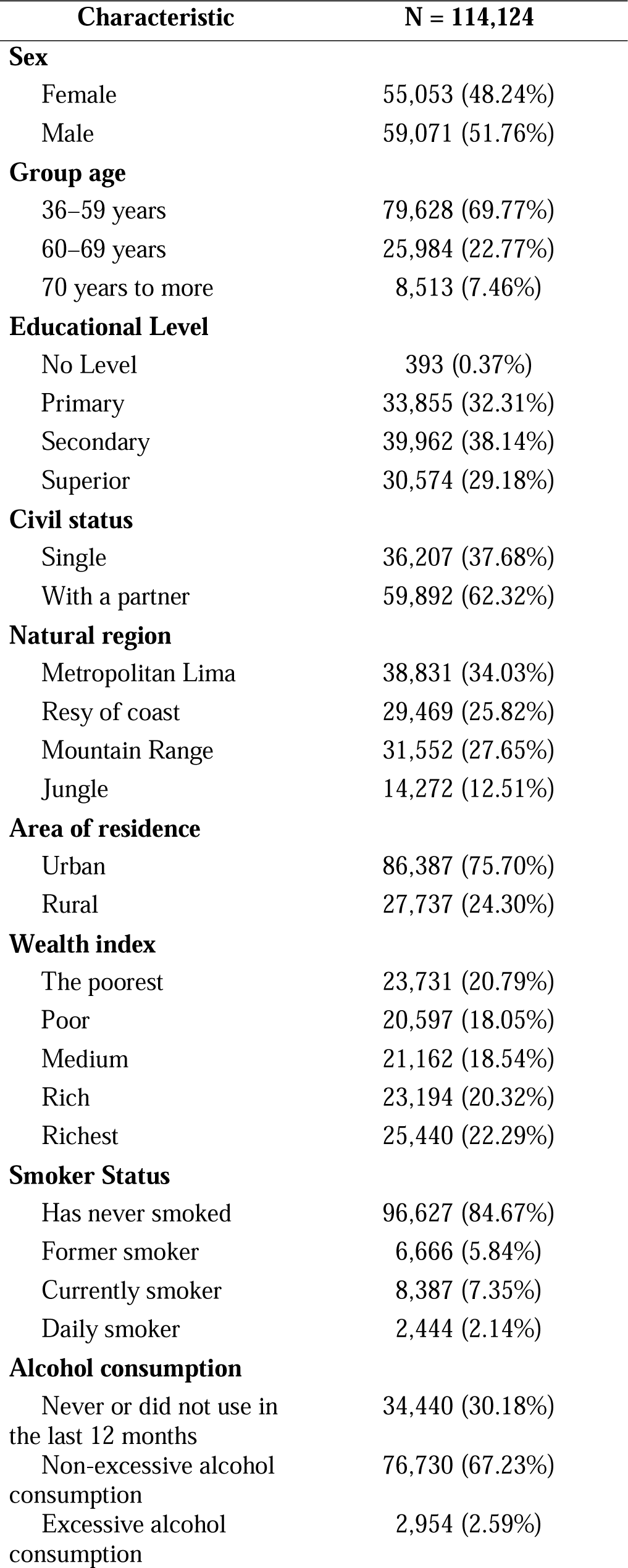
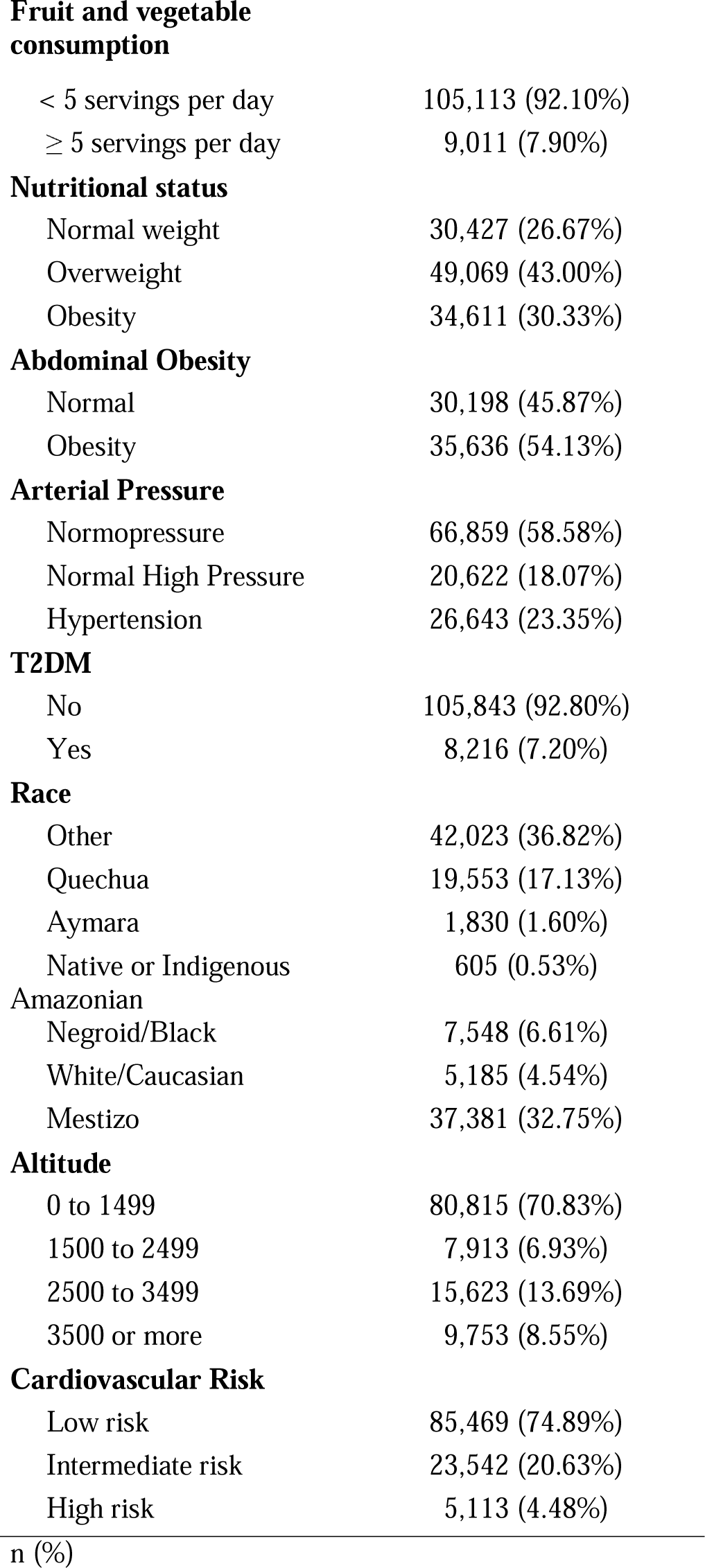
Descriptive characteristics of the study sample.

| Characteristic | N = 114,124 |
| --- | --- |
| **Sex** |  |
| Female | 55,053 (48.24%) |
| Male | 59,071 (51.76%) |
| **Group age** |  |
| 36–59 years | 79,628 (69.77%) |
| 60–69 years | 25,984 (22.77%) |
| 70 years to more | 8,513 (7.46%) |
| **Educational Level** |  |
| No Level | 393 (0.37%) |
| Primary | 33,855 (32.31%) |
| Secondary | 39,962 (38.14%) |
| Superior | 30,574 (29.18%) |
| **Civil status** |  |
| Single | 36,207 (37.68%) |
| With a partner | 59,892 (62.32%) |
| **Natural region** |  |
| Metropolitan Lima | 38,831 (34.03%) |
| Resy of coast | 29,469 (25.82%) |
| Mountain Range | 31,552 (27.65%) |
| Jungle | 14,272 (12.51%) |
| **Area of residence** |  |
| Urban | 86,387 (75.70%) |
| Rural | 27,737 (24.30%) |
| **Wealth index** |  |
| The poorest | 23,731 (20.79%) |
| Poor | 20,597 (18.05%) |
| Medium | 21,162 (18.54%) |
| Rich | 23,194 (20.32%) |
| Richest | 25,440 (22.29%) |
| **Smoker Status** |  |
| Has never smoked | 96,627 (84.67%) |
| Former smoker | 6,666 (5.84%) |
| Currently smoker | 8,387 (7.35%) |
| Daily smoker | 2,444 (2.14%) |
| **Alcohol consumption** |  |
| Never or did not use in the last 12 months | 34,440 (30.18%) |
| Non-excessive alcohol consumption | 76,730 (67.23%) |
| Excessive alcohol consumption | 2,954 (2.59%) |
| **Fruit and vegetable consumption** |  |
| < 5 servings per day | 105,113 (92.10%) |
| ≥ 5 servings per day | 9,011 (7.90%) |
| **Nutritional status** |  |
| Normal weight | 30,427 (26.67%) |
| Overweight | 49,069 (43.00%) |
| Obesity | 34,611 (30.33%) |
| **Abdominal Obesity** |  |
| Normal | 30,198 (45.87%) |
| Obesity | 35,636 (54.13%) |
| **Arterial Pressure** |  |
| Normopressure | 66,859 (58.58%) |
| Normal High Pressure | 20,622 (18.07%) |
| Hypertension | 26,643 (23.35%) |
| **T2DM** |  |
| No | 105,843 (92.80%) |
| Yes | 8,216 (7.20%) |
| **Race** |  |
| Other | 42,023 (36.82%) |
| Quechua | 19,553 (17.13%) |
| Aymara | 1,830 (1.60%) |
| Native or Indigenous Amazonian | 605 (0.53%) |
| Negroid/Black | 7,548 (6.61%) |
| White/Caucasian | 5,185 (4.54%) |
| Mestizo | 37,381 (32.75%) |
| **Altitude** |  |
| 0 to 1499 | 80,815 (70.83%) |
| 1500 to 2499 | 7,913 (6.93%) |
| 2500 to 3499 | 15,623 (13.69%) |
| 3500 or more | 9,753 (8.55%) |
| **Cardiovascular Risk** |  |
| Low risk | 85,469 (74.89%) |
| Intermediate risk | 23,542 (20.63%) |
| High risk | 5,113 (4.48%) |

**Table 2.**
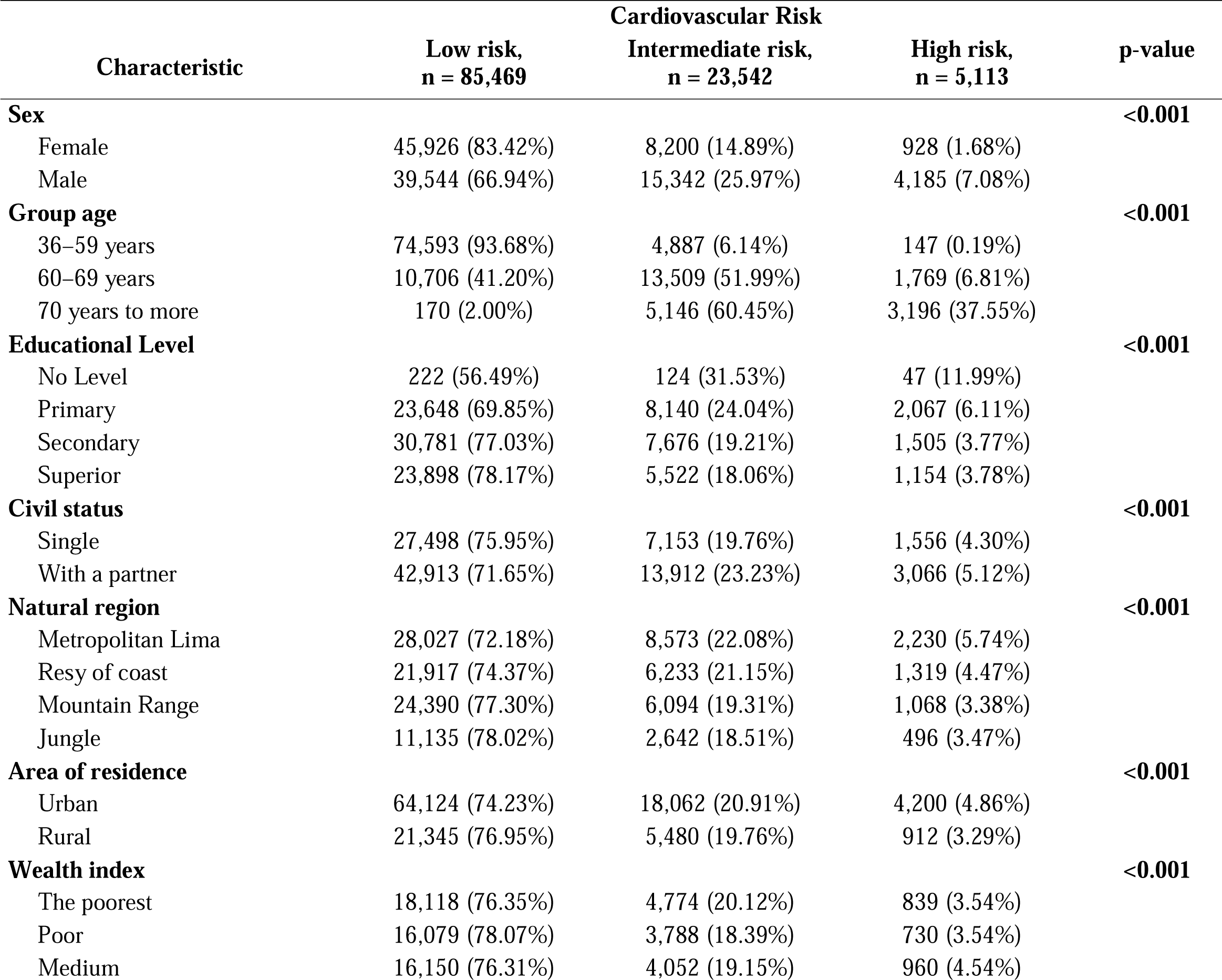
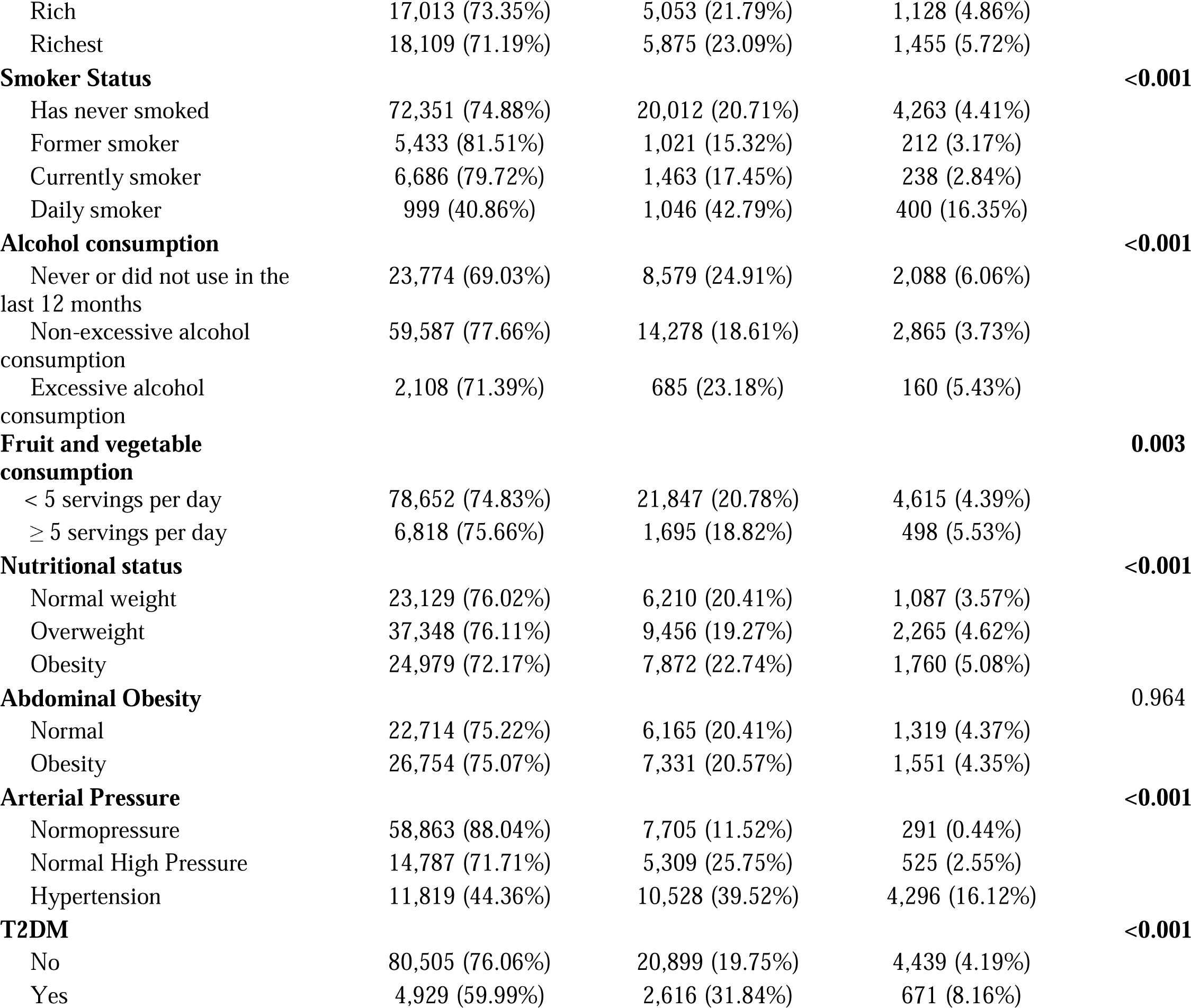
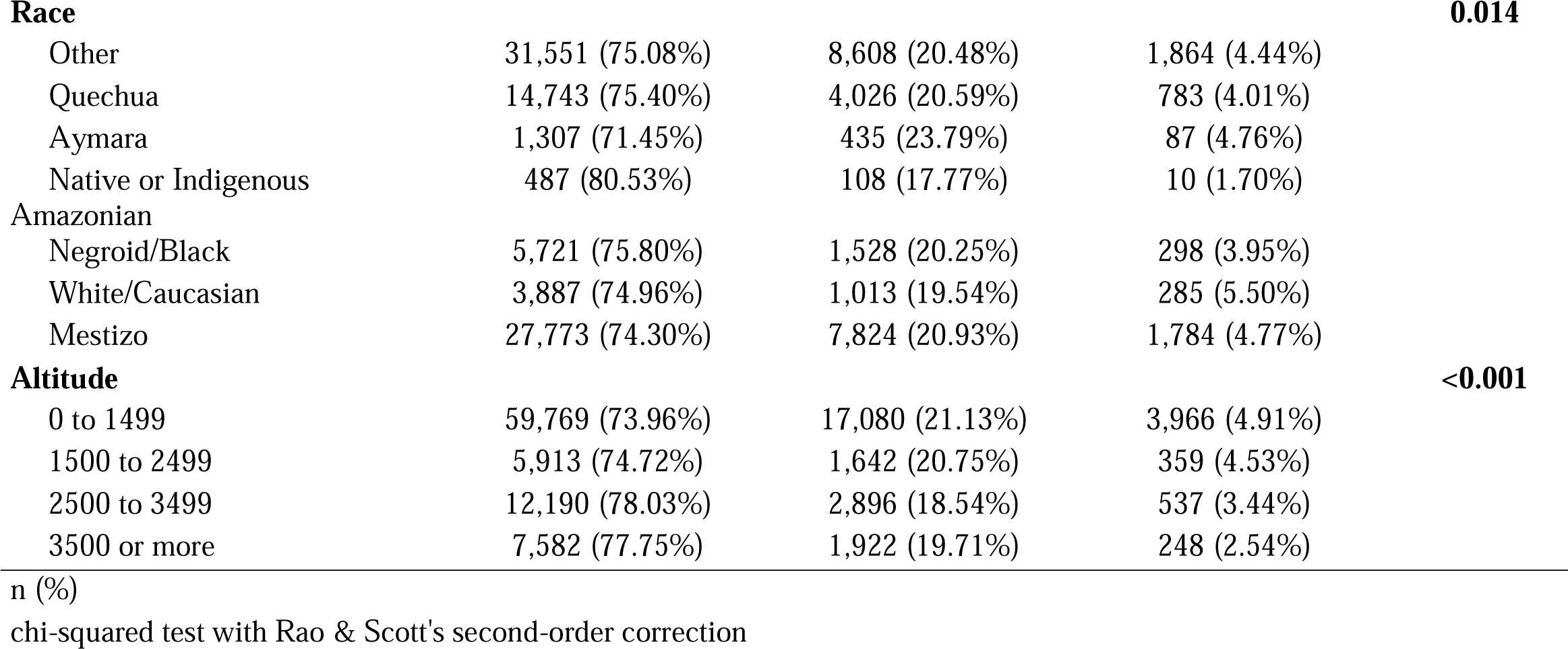
Bivariate analysis of the factors associated with CVR.

**Table 3.** Multivariable regression analysis of factors associated with CVR.

| Characteristic | aPR | 95% CI | p-value |
| --- | --- | --- | --- |
| **Sex** |  |  |  |
| Female | Ref. |  |  |
| Male | 2.36 | 2.26, 2.46 | <0.001 |
| **Group age** |  |  |  |
| 36–59 years | Ref. |  |  |
| 60–69 years | 7.56 | 7.22, 7.93 | <0.001 |
| 70 years to more | 11.3 | 10.7, 11.9 | <0.001 |
| **Educational Level** |  |  |  |
| No Level | Ref. |  |  |
| Primary | 0.81 | 0.66, 1.01 | 0.053 |
| Secondary | 0.78 | 0.63, 0.98 | **0.025** |
| Superior | 0.71 | 0.57, 0.89 | **0.002** |
| **Civil status** |  |  |  |
| Single | Ref. |  |  |
| With a partner | 1.03 | 0.99, 1.06 | 0.134 |
| **Natural region** |  |  |  |
| Metropolitan Lima | Ref. |  |  |
| Resy of coast | 1 | 0.96, 1.05 | 0.955 |
| Mountain Range | 0.97 | 0.84, 1.13 | 0.704 |
| Jungle | 0.99 | 0.92, 1.06 | 0.699 |
| **Area of residence** |  |  |  |
| Urban | Ref. |  |  |
| Rural | 1.01 | 0.94, 1.09 | 0.794 |
| **Wealth index** |  |  |  |
| The poorest | Ref. |  |  |
| Poor | 1.05 | 0.97, 1.14 | 0.212 |
| Medium | 1.04 | 0.96, 1.14 | 0.345 |
| Rich | 1.15 | 1.05, 1.26 | **0.002** |
| Richest | 1.13 | 1.03, 1.24 | **0.009** |
| **Smoker Status** |  |  |  |
| Has never smoked | Ref. |  |  |
| Former smoker | 0.93 | 0.85, 1.02 | 0.133 |
| Currently smoker | 0.98 | 0.91, 1.06 | 0.676 |
| Daily smoker | 1.91 | 1.75, 2.07 | <0.001 |
| **Alcohol consumption** |  |  |  |
| Never or did not use in the last 12 months | Ref. |  |  |
| Non-excessive alcohol consumption | 0.96 | 0.92, 1.00 | **0.031** |
| Excessive alcohol consumption | 1.02 | 0.92, 1.12 | 0.743 |
| **Fruit and vegetable consumption** |  |  |  |
| < 5 servings per day | Ref. |  |  |
| ≥ 5 servings per day | 0.99 | 0.93, 1.06 | 0.848 |
| **Nutritional status** |  |  |  |
| Normal weight | Ref. |  |  |
| Overweight | 1.04 | 0.99, 1.09 | 0.128 |
| Obesity | 1.18 | 1.11, 1.26 | **<0.001** |
| **Abdominal Obesity** |  |  |  |
| Normal | Ref. |  |  |
| Obesity | 1.13 | 1.07, 1.18 | **<0.001** |
| **Arterial Pressure** |  |  |  |
| Normopressure | Ref. |  |  |
| Normal High Pressure | 1.72 | 1.64, 1.81 | **<0.001** |
| Hypertension | 2.5 | 2.40, 2.61 | **<0.001** |
| **T2DM** |  |  |  |
| No | Ref. |  |  |
| Yes | 1.07 | 1.02, 1.13 | **0.009** |
| **Race** |  |  |  |
| Other | Ref. |  |  |
| Quechua | 0.97 | 0.90, 1.05 | 0.425 |
| Aymara | 0.97 | 0.86, 1.11 | 0.698 |
| Native or Indigenous Amazonian | 1.11 | 0.84, 1.43 | 0.448 |
| Negroid/Black | 0.99 | 0.90, 1.08 | 0.755 |
| White/Caucasian | 0.98 | 0.89, 1.07 | 0.605 |
| Mestizo | 0.97 | 0.91, 1.04 | 0.411 |
| **Altitude** |  |  |  |
| 0 to 1499 | Ref. |  |  |
| 1500 to 2499 | 1.03 | 0.90, 1.17 | 0.685 |
| 2500 to 3499 | 1.01 | 0.87, 1.18 | 0.865 |
| 3500 or more | 1.04 | 0.88, 1.23 | 0.638 |

Each factor has been independently adjusted for Sex, Group age, Educational Level, Civil status, Natural region, Area of residence, Wealth index, Smoker Status, Alcohol consumption, Fruit and vegetable consumption, Nutritional status, Abdominal Obesity, Arterial Pressure, T2DM, Race and Altitude
aPR: Prevalence ratio. CI 95%: Confidence interval at 95%
Source:
self-made

In terms of smoking status, 9.49% are current or daily smokers. Meanwhile, excessive alcohol consumers reach 2.59%. The consumption of fruits and vegetables is presented as more than five daily portions in only 7.90%. Additionally, those presenting with global and abdominal obesity were 30.33% and 54.13%, respectively. The prevalence of hypertension was 23.35%, and T2DM was 7.20%.

### Prevalence of CVR

The prevalence of low and high CVR was 74.89% and 4.48%, respectively. The graph shows the global CVR trends from 2014 to 2022. It is divided into two risk levels: moderate and high.

From 2014 to 2018, the percentage of moderate risk has shown an upward trend, starting at 18.54% in 2014 and reaching a peak of 25.08% in 2018. On the other hand, high risk has fluctuated more, starting at 4.78% in 2014, decreasing to 3.66% in 2015, and gradually increasing each year to 5.64% in 2017. Subsequently, it dropped slightly to 5.31% in 2018.

From 2019, a decrease in moderate risk is observed, falling to 18.9% and then varying slightly until reaching 19.68% in 2022. High risk shows a more consistent downward trend from 2019, from 4.25% to 3.82% in 2022.

### Factors Associated with CVR

The regression analysis identified several factors with a significant association in the adjusted prevalence ratio. Regarding gender, it was found that men have an increased risk compared to women. In terms of age, individuals between 60 and 69 years and those 70 years or older show a considerably higher risk than the reference group of 36 to 59 years.

Those with secondary and higher education have a lower risk than those without no education. In addition, wealth correlates with a higher risk; individuals classified as rich and most prosperous have a higher risk than the poorest group. Smoking habit is another significant factor, with daily smokers presenting a higher risk compared to those who have never smoked. Obesity, both general and abdominal, is associated with an increase in risk compared to those of average weight and without abdominal obesity, respectively. Meanwhile, health conditions such as high blood pressure and T2DM also showed a significant association, with a higher risk for people with these conditions compared to normotensive and non-diabetic individuals.

## Discussion

### Prevalence of CVR in Peru

While the graph depicts that the bulk of the inhabitants are generally situated within the moderate risk grouping in contrast to high risk, it simultaneously suggests that a sizeable proportion still faces significant vulnerabilities. While annual fluctuations have occurred, the share of people facing elevated danger has waned through the years. This trend may signify worldwide progress in handling the risk of cardiovascular troubles.

The results indicate a global prevalence of low and high CVR of 74.89% and 4.48%, respectively, with significant variations in trends from 2014 to 2022. The latest Global Burden of Disease report parallels these figures by demonstrating that deaths from cardiovascular diseases have risen from 12.4 million in 1990 to 19.8 million in 2022, attributable to population and demographic changes as well as metabolic, environmental, and behavioral risks that could have been avoided through prevention. Furthermore, the majority share of the worldwide burden of cardiovascular diseases has been shoulder by nations having a low or moderate income, especially within locales such as Oceania, Eastern, and Central Europe, as well as East and South Asia. These findings emphasize the pressing need for worldwide public health tactics to address and handle CVR proactively, specifically within underserved communities (15).

A systematic review and meta-analysis regarding Latin America published by Cagna-Castillo et al. (16) was examined in greater detail and identified reliable epidemiological evidence of stroke in Latin America and the Caribbean. The study determined that the general occurrence of stroke among participants was approximately 32 individuals out of every 1000, with the prevalence found to be remarkably similar for both sexes at around 21 and 20 people per 1000 for males and females, respectively. The overall incidence of stroke was 255 per-100,000 person-years, with the rate among males noticeably higher than females as males experienced 261 cases per 100,000 compared to females only suffering 217 per 100,000. These findings underscore the importance of understanding the frequency and rate of occurrence of stroke within the local area.

A recent CDC study reveals that, despite advances in CVD management, minority, disadvantaged, and underserved populations continue to experience significant health disparities exacerbated during the COVID-19 pandemic. Research has also underscored the necessity of tackling long-established CVD danger elements, like using tobacco and hypertension, through well-timed interventions and dissection of the social determinants influencing health outcomes. Furthermore, optimizing healthy behaviors and reducing the development and progression of CVD through a focus on prevention strategies underscores the necessity for evidence-based approaches to confront inequities in its prevention and management, as previously emphasized (17,18).

### Factors Associated with CVR

Our examination into the connection between gender and cardiovascular hazard uncovers meaningful divergences in how such sicknesses as ischemic heart illness and heart breakdown arise and conclude. While biological factors like genes and sex hormones contribute to variances, as does the societal construct of gender, these aspects have dissimilar impacts on how each sex frequently behaves and is treated in cultures worldwide. At the same time, observations have noted that variances in sex may often manifest opposing impacts on clinical presentations and consequences of cardiovascular ailments in comparison to gender-linked elements. These differences exist from birth and are determined by purely biological mechanisms. Additionally, the predominantly male leadership and workforce in clinical cardiology may be disadvantageous for female patients. Risk factors for cardiovascular diseases related to the female sex include complications during pregnancy, therapy for breast cancer, autoimmune and rheumatic diseases, depression, and stress related to home life. Despite more favorable biology, gender-related factors worsen outcomes in women with coronary disease or heart failure compared to men (19–21).

Our findings demonstrate a notable association between the hazard of cardiovascular ailments and the advancing years of those in the group we evaluated. We discovered that those within the 60-69 age range, as well as individuals 70 or older, face an elevated risk of CVR compared to the benchmark group aged 36 to 59 years old. The existing literature corroborates this observation by indicating that advancing age contributes significantly to cardiovascular deterioration and the ensuing heightened risk of cardiovascular diseases amongst the elderly. With advancing age, both sexes confront growing chances of atherosclerosis, strokes, and heart attacks, illnesses that compose the more extensive assortment of cardiovascular diseases. Older patients, especially those over seventy-five, face heightened vulnerability to coronary artery disease due to the considerable impacts of advancing age on atherosclerosis progression and worse anticipated outcomes. Indeed, CAD remains a primary contributor to mortality and poor health outcomes within this group, as evidenced by previous research. By 2030, projections indicate that a fifth of Americans will have entered retirement, with over 20 percent of the population comprising those aged 65 years or more, as the AHA anticipates a sizable aging of citizens over the next decade. In this age group, cardiovascular diseases are projected to account for nearly two in every five mortalities, maintaining their position as the primary producer of demise, according to current statistics. Additionally, those within the 60-80 age range tend to exhibit cardiovascular issues at rates of around 75-78% on average. In contrast, for patients over 80, the likelihood of such diseases rises to exceed 85% (22).

Our results indicate an inverse correlation between educational level and CVR. We observe that those with secondary and higher education have a lower risk of CVR than those without formal education. This finding aligns with past work demonstrating that higher levels of educational achievement tend to be associated with a lowered prevalence of specific cardiovascular disease risk factors, such as T2DM, hypertension, and abnormal lipid profiles, as demonstrated through a collection of studies illustrating an indirect linkage between these variables (23). Additional studies propose that the level of education attained notably impacts the likelihood of developing heart conditions, emphasizing how acquiring knowledge represents a crucial societal influencer on individual well-being, as previously reported (24). Furthermore, research exploring the connection between educational attainment and cardiovascular disease risk across the lifespan has evaluated education as an essential determinant for anticipating cardiovascular disease hazards based on existing evidence (25). Studies have additionally noted how the degree of formal education that one attains can impact the projected outcomes regarding cardiovascular illnesses, such as stroke, owing to dissimilarities in the regularity of conventional risk elements between groups with divergent levels of academic accomplishment (26).

Surprisingly, though conventional wisdom suggests otherwise, our findings reveal a tendency for prosperity and charitable acts to accompany one another frequently. Specifically, we observe that wealthy and affluent individuals present a higher risk of CVR than the poorest group. A study published by Machado S et al. (27) found that changes in wealth mobility are associated with long-term cardiovascular health outcomes. Specifically, negative wealth mobility is associated with a higher risk of cardiovascular events, while positive changes in wealth are associated with a lower risk. In addition, it is mentioned that the commonwealth is a dynamically changing risk factor that can influence a person’s cardiovascular health status throughout life. Additionally, decreases in wealth are associated with more stress, fewer healthy behaviors, and less leisure time, all of which are associated with worse cardiovascular health.

Our findings highlight that smoking, especially daily smoking, is a significant factor in increasing CVR. The data demonstrates that those who smoke daily face a higher risk of cardiovascular issues than people who have avoided tobacco throughout their lifetime. A manuscript published by Shields and Wilkins (28) underscores the notable danger that smoking poses as a risk factor for heart disease by finding those who smoked daily during the follow-up period faced a 60% elevated risk of incident heart disease relative to individuals who had never smoked daily. Furthermore, the International CLARIFY registry investigation of patients with stable coronary artery disease demonstrated that both current and former tobacco consumers faced an elevated likelihood of all-cause and cardiovascular mortality when placed next to those who had never smoked (29). Additional research leveraging information from the National Health and Nutrition Examination Survey similarly demonstrated a considerably heightened probability of all-cause mortality as well as mortality stemming from cardiovascular ailments and cancers among individuals smoking daily (30).

Our findings suggest that when considering general and abdominal obesity, risks to cardiovascular health rise in contrast with persons maintaining an average weight or who lack excess weight gain. The global surge in obesity, having steadily grown since the 1980s, has a direct hand in heightening vulnerabilities to heart troubles like abnormal lipid levels, T2DM, high blood pressure, and sleep issues. Additionally, the development of cardiovascular diseases and mortality resulting from such conditions can be independently attributed to obesity aside from other CVR factors (31). Recent research underscores abdominal obesity, quantified via waistline, as a distinguishing sign of cardiovascular ailment exposure regardless of body mass gauge, redrawing attention to waist circumference as a risk evaluation with meaning separate from BMI. Furthermore, research has discovered that measurements of abdominal obesity relating to those of both sexes who maintain a standard BMI are connected to an elevated potential for encountering determinants signaling cardiovascular conditions (32).

Our results show a significant association between high blood pressure and T2DM with an increase in CVR compared to normotensive and non-diabetic individuals. There is a well-established correlation between hypertension and T2DM. Having hypertension appears to increase the risk of T2DM, and having T2DM increases the risk of hypertension. Both conditions are aspects of the underlying syndrome, including obesity and heart disease. The combination of hypertension and T2DM is particularly lethal and can significantly increase the risk of suffering a heart attack or stroke. Epidemiological studies indicate that there is a very high incidence of hypertension and associated cardiovascular disease in patients with T2DM. Additionally, hypertension and T2DM are common comorbidities, with hypertension being twice as frequent in diabetic patients compared to non-diabetic individuals. A large group of prospective studies has associated T2DM with an increased risk of hypertension.

### Public Health Importance

The relevance of our study transcends borders, offering valuable insights for public health at both national and international levels. The findings lay crucial groundwork for crafting more impactful health policies and targeted prevention tactics customized for various demographic groups. This study’s identification of factors including age, education, wealth, smoking, obesity, hypertension, and T2DM underscores opportunities for interventions to most positively impact those facing higher risks from specified conditions. These data are essential for designing specific prevention and management programs that can be implemented locally and globally, considering cultural and socioeconomic variations.

Moreover, by providing evidence on how various socioeconomic and lifestyle factors influence CVR, this study can guide policymakers and health professionals in prioritizing resources and efforts. By applying these understandings to public health policies, one may potentially decrease the worldwide weight of CVD, enhancing the standard of living and diminishing well-being disparities experienced globally. While the interplay of risk factors demands a comprehensive strategy, these findings might spur governments and international bodies to reaffirm their dedication to communal cardiac welfare, acknowledging how promoting overall wellness depends on addressing linked dangers through an inclusive approach.

### Choice of CVR Calculator

The CVR calculators are pivotal tools in preventing and managing cardiovascular diseases. The WHO’s risk calculator in our study offers the advantage of not requiring laboratory tests, making it particularly suitable for global application. This feature is especially beneficial in contexts where laboratory data are unavailable, as in the National Survey of Demography and Family Health (ENDES) case or in populations lacking access to frequent laboratory evaluations. Although the WHO’s risk calculator has been evaluated in 21 countries globally, it’s notable that it includes minimal data from Latin American countries (33,34). However, a significant limitation of this calculator is its exclusion of key biomarkers like cholesterol and triglycerides, which are crucial in assessing CVR (35).

In contrast, the Framingham and American Heart Association (AHA) calculators include laboratory analyses in their assessments (36,37). These calculators offer a more detailed risk estimation but rely on information that may not always be available in public health surveys or low-resource settings.

A significant limitation of existing CVR calculators is their lack of specific adaptation to Latin American populations. Most calculators have been developed and validated in high-income country populations. They may not accurately reflect the risk profiles of Latin American populations, where factors like genetic differences, lifestyles, and socioeconomic conditions significantly influence CVR (38,39).

To improve CVR assessment in Latin America, it is crucial to develop and validate CVR calculators tailored to the specific characteristics of these populations (40). This involves gathering regional epidemiological data and incorporating local risk factors into the calculators. Additionally, fostering research in biomarkers and region-specific risk factors, which could be integrated into future versions of CVR calculators, would be beneficial. Collaboration among public health institutions, researchers, and international organizations is critical to achieving these goals.

In conclusion, while the WHO CVR calculator provides a valuable tool for global risk assessment, its limitations highlight the need for more region-specific calculators, particularly for Latin American populations. Developing such tailored tools would significantly enhance the accuracy of CVR assessments and the effectiveness of corresponding health interventions in these regions.

### Study Limitations

As the study is cross-sectional in design, it does not allow for causal inferences. Although associations between the studied variables and CVR can be identified, cause-and-effect relationships cannot be established. Furthermore, the study focused on individuals aged 40 to 75 years, limiting the generalizability of the results to the entire Peruvian population. Additionally, exclusions based on the lack of interest variables could introduce biases in the results.

Although practical, the WHO calculator does not include laboratory analysis, which may limit the precision in assessing CVR. This fact might underestimate or overestimate CVR in some individuals. Also, although parameters were established to ensure practical blood pressure measurements, the study relied on self-reported data for other risk factors like smoking and T2DM, which can have a particular recall bias.

### Conclusions

The study revealed a significant prevalence of CVR in the studied population. The critical importance of addressing these prevalent factors in public health policies and strategies for preventing and managing cardiovascular diseases is strongly highlighted. Age, educational attainment, wealth, daily smoking, obesity measures, and health issues, including hypertension and T2DM, demonstrated robust links with heightened cardiovascular danger, according to findings.

Consequently, implementing targeted public health initiatives focused on preventing and treating CVR factors common in older populations seems urgently warranted, considering their elevated susceptibility. Promoting cardiovascular health education across all levels of education is crucial to raise awareness and better prevent cardiovascular diseases by improving understanding from a young age.

Furthermore, interventions addressing lifestyles and stress in higher-wealth populations should be developed, considering their association with increased CVR. Intensifying campaigns for tobacco cessation and obesity control at the community and national levels are essential, given their direct relationship with increased CVR. There is a pressing need to strengthen the holistic handling of hypertension and T2DM through timely identification and suitable intervention to circumvent cardiovascular issues. Promoting the creation and validation of CVR calculators adapted to Latin American populations, including region-specific risk factors, is essential.

## Acknowledgments

A special thanks to the members of Instituto de Investigación de Enfermedades Tropicales, Universidad Nacional Toribio Rodríguez de Mendoza de Amazonas (UNTRM), Amazonas, Perú who provided valuable comments during the preparation of this study.

## Informed consent

It was not necessary to obtain informed consent in this Study.

## Data availability

The data supporting the findings of this study can be accessed by the original research paper at the follow link: https://proyectos.inei.gob.pe/microdatos/

## Author contributions

**Víctor Juan Vera-Ponce:** Methodology, Data Analysis, Writing – Review & Editing.

**Fiorella E. Zuzunaga-Montoya:** Conceptualization, Data Analysis, Writing – Review & Editing.

**Joan A. Loayza-Castro:** Data Analysis, Methodology, Validation, Writing – Original Draft.

**Luisa Erika Milagros Vásquez Romero:** Data Analysis, Methodology, Validation, Writing – Original Draft.

**Mario J. Valladares-Garrido:** Visualization, Validation, Project Administration, Supervision, Methodology, Writing – Review & Editing.

**Carmen Inés Gutierrez De Carrillo:** Supervision, Methodology, Funding Acquisition, Writing – Review & Editing.

---

**Figure 1.**
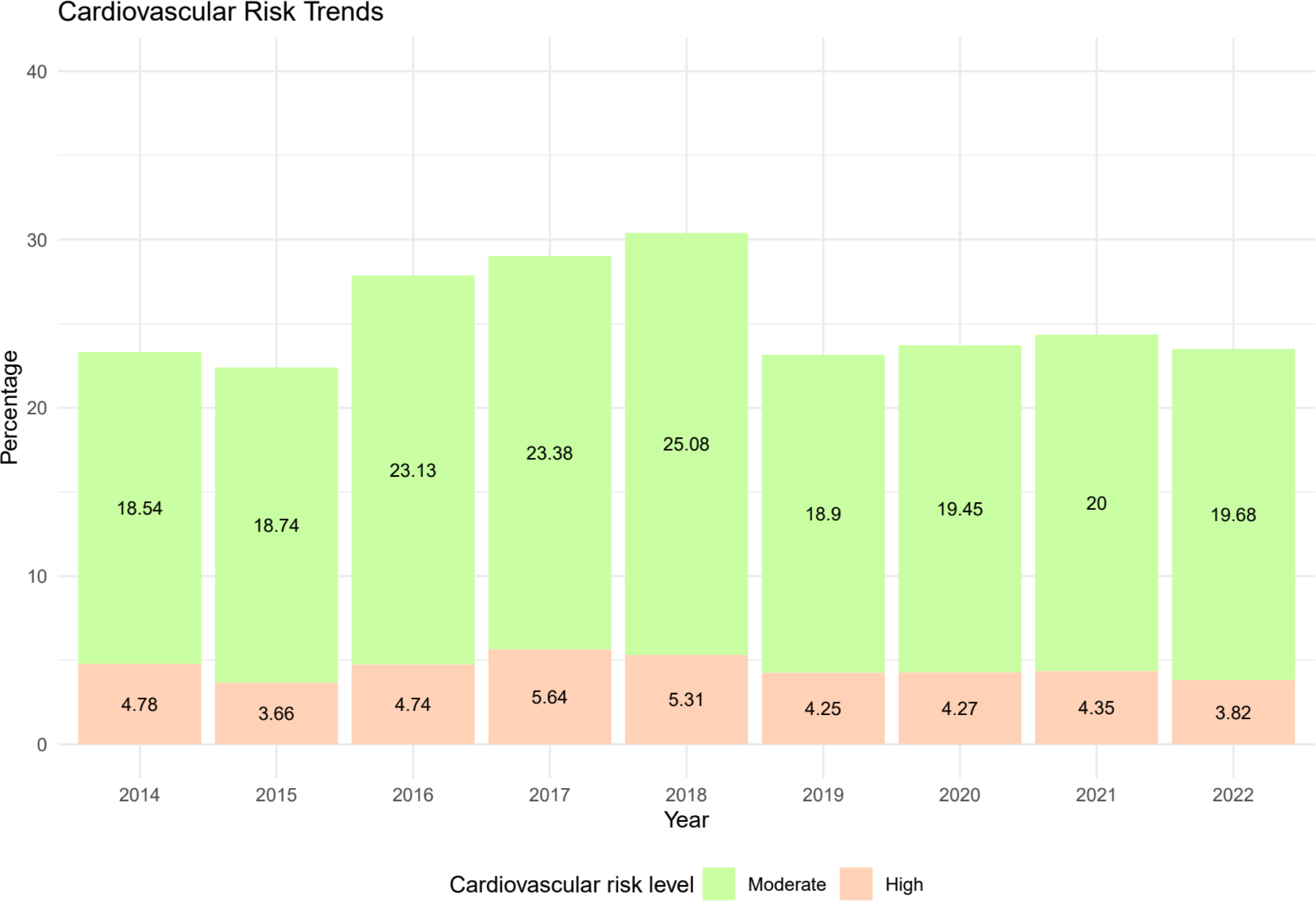
Trend graph of Cardiovascular Risk in Peru (2014 – 2022).

